# Prioritizing areas for multisectoral interventions (PAMIs) for cholera control in Cameroon

**DOI:** 10.1101/2025.01.03.25319978

**Authors:** Armelle Ngomba, Linda Esso, Nicole Fouda Mbarga, Ingrid Kenko, Eric Defo, Nadia Mandeng, Theodore A. Tonye, Patricia Mendjime, Chanceline Bilounga, Loic Choupo, Emmanuel Douba, Georges A Etoundi

**Author notes:** **Corresponding author:** Dr Nicole Fouda Mbarga, World Health Organisation (WHO) Cameroon, Yaounde, Cameroon.

## Abstract

**Introduction:** The Global Task Force for Cholera Control (GTFCC) developed a Global Roadmap for Ending Cholera by 2030 emphasizing the need for targeted multisectoral interventions in priority areas. In 2023 for the first time in Cameroon, identification of PAMIs for cholera control was conducted, following the 2023 GTFCC guidelines. We hereby describe experience of PAMI’s identification in a cholera high-burden country.

**Methods:** In August 2023, a multisectoral technical team was set up, under the leadership of the Ministry of health to conduct identification of *PAMIs.* Stakeholders were briefed on GTFCC guidelines. A retrospective (January 2016-September 2023) descriptive study was conducted in all the 10 regions of Cameroon. Data were collected from DHIS-2, national cholera line lists, situation reports, databases from the Centre Pasteur of Cameroon and the National Public Health Laboratory. These datasets were entered in the GTFCC tool for automated analysis. An initial list of PAMIs was identified based on a priority index using four indicators: cholera incidence, mortality, persistence, and positivity. In December 2023, we conducted a ***multisectoral validation workshop****, during which the threshold of 9 was set* for PAMIs’ prioritization *and a* second prioritization done using vulnerability factors.

**Results:** Overall, 48 health districts (25% of all health districts) were identified as PAMIs, 35 PAMIs using the priority index and 13 based on vulnerability factors. About 11 488 089 people (41% of the country’s population) live in these PAMIs from which 93% of cholera cases were reported over the past 8 years. The Centre, Littoral, South-West, and the Far North regions are home to 66% of PAMIs.

**Conclusion:** Identification of PAMI’s was successfully conducted in Cameroon and provided scientific evidence for decision-making to level up cholera preparedness and prevention. The ownership and leadership of governmental main stakeholders were critical. Data availability facilitated this exercise.

**Key questions:** *What is already known on this topic:* Ending cholera requires implementation of targeted interventions in high-priority areas for maximum impact. Identification of these areas (PAMIs) is the first step towards cholera elimination.

*What this study adds:* Priority areas for targeted cholera control interventions were identified and categorized in Cameroon for cholera control. PAMIs represent 25% of health districts in Cameroon with a targeted population representing 41% of the whole country. The study describes lessons learnt and challenges of using GTFCC guides for PAMIs identification, which is useful information for other countries.

*How this study might affect research, practice or policy:* Our findings will inform decision-making for cholera control in Cameroon and in other countries through elaboration, implementation and monitoring and evaluation of a national cholera control budgeted plan with specific key long-term interventions to be conducted where needed.

## Introduction

Cholera is an infectious acute watery diarrheal disease resulting from ingestion of contaminated water or food with toxigenic form of Vibrio cholerae O1 and O139^1,2^. Cholera trends worldwide portray a picture of a threatening global health challenge, a marker of inequity and poverty^2^ with the highest burden in Eastern Mediterranean region and Africa^3^. The main determinants of cholera are insufficient access to potable water, basic sanitation, and hygiene (WASH) facilities which are aggravated by environmental disasters such as floods or droughts, population displacement, conflicts, and poor access to healthcare^4–6^.

Located at the heart of Central Africa, Cameroon is particularly vulnerable to cholera and the country has faced several outbreaks since 1971^7^. In Cameroon, cholera occurs during the rainy season in southern regions and during the dry seasons in the northern site of the country, but the rainy season has been identified as a risk factor for cholera in the country (OR 0.60, 95% CI 0.45-0.78)^7^. Cameroon has a vast maritime border extending from the Bight of Biafra, part of the Gulf of Guinea and the Atlantic Ocean^8^. Cameroon is bordered by Chad in the Northeast, the Central African Republic in the East, and Equatorial Guinea, Gabon and the Republic of the Congo in the South and by Nigeria in the West and North. Cholera remains a significant public health problem in Nigeria especially in Borno and Adamawa states which borders Cameroon, increasing the risks of cross-border transmission^9^. Additionally, Cameroon is a lower-middle-income country with a population of over 27.9 million (2022)^10^. According to the joint monitoring report (JMP report), Cameroon is one of the countries whose national situation is critical with 30% of populations having no access to drinking water, only 10.9% of households using drinking water techniques appropriate water treatment and 36% of the population has access to basic hygiene^11^. Moreover, the security context in some regions exacerbates these needs. According to OCHA, Cameroon bears the burden of three protracted crises (the conflict of the Lake Chad Basin, the crisis in the North-West and South-West regions, the influx of Central African refugees) epidemics and climate shocks^12^. In Cameroon, nearly half a million people are internally displaced and the country hosts almost a half a million refugees and asylum seekers^12^.

From 2018 to 2023, a persistence of cholera cases was observed in Cameroon with a peak in 2022 during which 13,420 cases and 251 deaths (TL=1.9%) were recorded^13^. This persistent public health threat prompted the development of a National Cholera Plan (NCP) from 2023 in Cameroon. According to GTFCC guidance, there are 4 main steps for the development of the NCP: an inception phase during which countries make a political commitment and identify cholera PAMIs, the development phase, an implementation phase and the monitoring and reporting phase^14,15^. A PAMI is a “geographically limited area where environmental, cultural and/or socioeconomic conditions facilitate the transmission of the disease, and where cholera persists or reappears regularly”^16^. Cholera preparedness and response is limited by resources^3^, thus, identifying, and prioritizing areas most at risk (PAMIs) is an imperative for cholera control.

In 2023, the GTFCC published new guidelines for the identification of priority areas for multisectoral interventions (PAMIs), based on cholera incidence, mortality, persistence, and positivity and optional vulnerability factors ^16^. In accordance with GTFCC recommendations, in 2023 Cameroon started the process of identifying PAMIs at the district level. The current study aimed to describe the process of identification and classification of priority areas for multisectoral interventions (PAMIs) for cholera control in Cameroon at the district level using the GTFCC 2023 guidelines for prioritization of PAMIs.

## Material and methods

### Study design and site

To achieve our goal of identifying PAMIs, we conducted retrospective cross-sectional descriptive study of the health map of Cameroon, which includes the 10 regions and 203 health districts, divided into 1819 health areas. Datasets were prepared with district level data covering 8 years from January 2016 to September 2023. A national multisectoral technical team was set up to conduct this exercise under the leadership of the Ministry of health with the support of partners. A technical briefing was conducted by the WHO using GTFCC guidelines.

### Cholera case definition

A suspected cholera case was defined as an individual with one of the two following conditions^18^:

In regions where a cholera epidemic has not been declared: any patient aged two years and older with acute watery diarrhea and severe dehydration OR who dies from acute watery diarrhea. In regions where a cholera epidemic has been declared, any person presenting with or dying from acute watery diarrhoea.

Furthermore, in health facilities and at the community level, a suspected cholera case (often referred to as the community case definition) was defined as any person two years of age or more with profuse AWD and/or vomiting^18^. While a confirmed case refers to a suspected case of Vibrio cholerae strain O1 or O139 confirmed by culture or gene amplification and, in countries where cholera is not present or has been eliminated, Vibrio cholerae strain O1 or O139 is toxigenic.

### Cholera data sources

Data sources were the DHIS-2, the national cholera line lists, the national and regional situation reports, the Centre Pasteur of Cameroon, the National Public Health Laboratory Databases. The QGIS software (version 3.34) was used to map PAMIs. The cholera line lists for the period week 01 2016 to week 53 2023 were provided by the Direction of the Fight against Epidemics and Pandemics (DLMEP) of the Ministry of Health in Cameroon.

### Population data

The Cameroon population data projections per district for the years 2016-2023 were obtained from Health Information Unit of the Ministry of Health in Cameroon.

### Geographic information system (GIS) data

The mapping of identified PAMI was done using the original GIS file layers (shapefile). These included shapefiles for regional administrative units (10 regions) and health district-level administrative units (203 health districts). Additionally, we used original shapefiles specific to neighboring countries. this file layers are updated annually and made available to teams by the Health Information Unit of the Ministry of Health in Cameroon.

### Vulnerability data

Dataset for cholera vulnerability assessment was generated using a KoboCollect tool filled at the regional and district level. Taking into consideration vulnerability factors for cholera is essential to identify and prioritize hotspots (PAMIs).

The main vulnerability factors assessed were WASH conditions, population movements, insecurity, accessibility, poverty, risk of cross-border transmission (neighboring countries) etc. These factors are key in identifying high risk areas and directing cholera prevention and control actions appropriately.

### Data analysis

Two datasets were inserted in the GTFCC 2023 Excel tool for automated analysis: an epidemiological dataset (cholera data and population) and vulnerability data. Quantitative epidemiological variables of interest such as cholera incidence, mortality, persistence, and positivity were calculated for each health district using the tool. Scores were assigned to each of these indicators and a priority index was calculated generated by summing up these scores. Based on this, districts were ranked according to their priority index, thereby determining priority levels for cholera interventions.

### Multisectoral validation and prioritization of PAMIs

A multi-sectoral workshop was organized under the lead of the Prime Minister’s services. This workshop brought together heads of different government departments as well as technical and financial partners. The results of PAMIs identification and prioritization using epidemiological data and vulnerability factors were presented and discussed during this workshop, allowing consensual validation of priority areas for intervention against cholera. The country set up a threshold of 9 for prioritization of PAMIs. At the end of this process, a final list of PAMIs was established, including health districts identified as having a high priority index as well as those presenting significant vulnerability factors.

### Patient and public involvement

The public represented by the civil society was involved in the multisectoral stakeholder consultation and validation of these results.

## Results

### Database description

Between 2016 and 2023, Cameroon notified 24,813 cholera suspected cases in 09 regions with a persistence rate of 51.2% across 213 epidemiological weeks out of 416 weeks. Among these cases, 14,036 were tested for *vibrio cholerae* confirmation by rapid diagnostic test (RDT) or culture, with a positivity rate of 67.13% (Table 1). A total of 680 deaths were recorded during the same period, resulting in a case fatality rate of 2.74%. Over the same period, we observed an incidence and mortality rate of 11.52 and 0.32 per 100,000 inhabitants, respectively (Table 1).

**Table 1:**
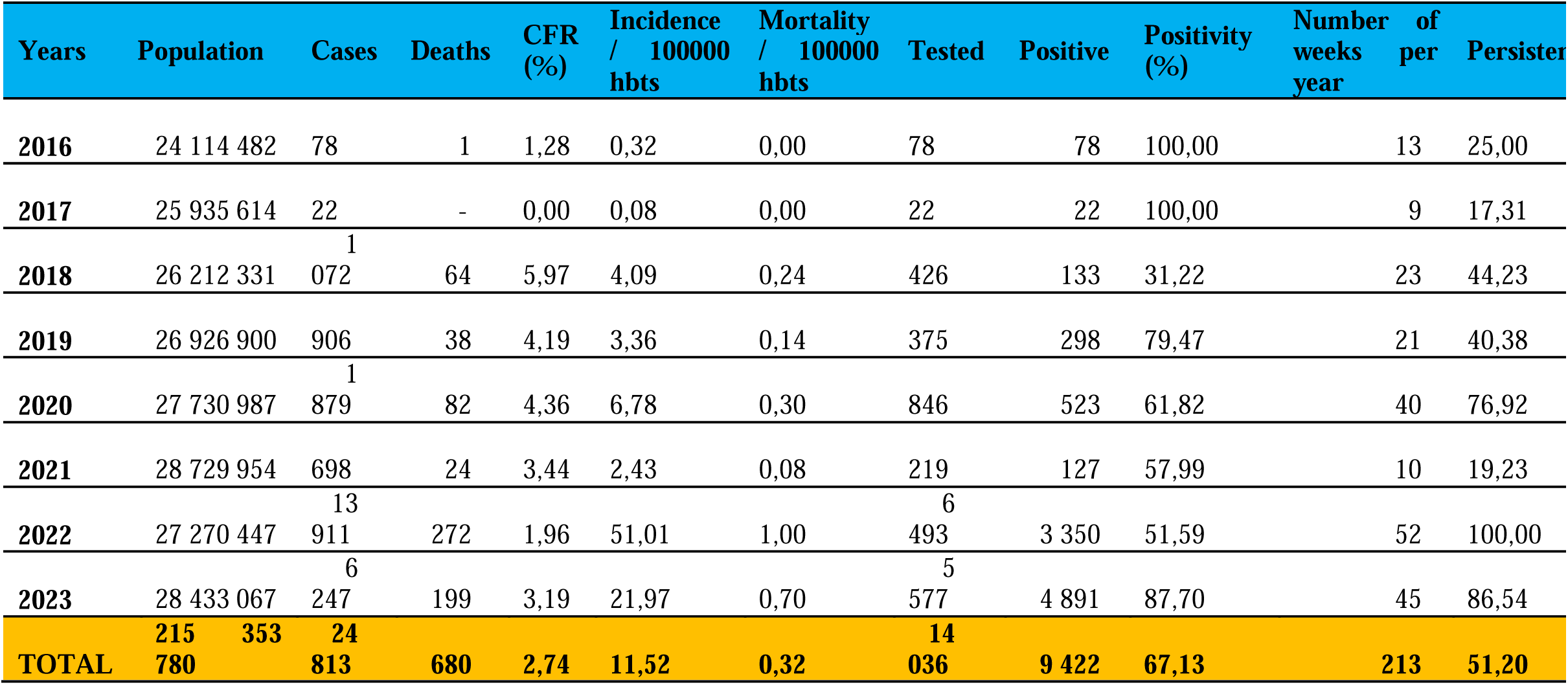
Summary of cholera epidemiology in Cameroon (2016-2023)

### Evolution and Mapping of Cholera Cases in Cameroon between 2016 and 2023

Between 2016 and 2023, the region of Adamawa did not report any confirmed cholera cases. In 2016, six regions were affected (Centre, East, Far-North, Littoral, North, and South), with the Centre reporting the most cases (35 without deaths, CFR 0%) and Littoral the fewest cases (1 case with death, CFR 100%). In 2017, four regions were affected (Centre, Littoral, North, and West), with Littoral having the highest incidence (18 with no death, CFR 0%) and North and West regions the fewest cases (1 case each, without deaths, CFR 0%). In 2018, five regions were affected (Centre, Far-North, Littoral, North, and South-West), and the North reported most of the cases (637 with 41 deaths, CFR 6.44%) and South-West the least number of cases (1 case without death, CFR 0%). In 2019, two regions were affected (North and South-West), with the North reporting most cases (536 with 22 deaths, CFR 4.10%) and South-West the fewest cases (370 cases with 16 deaths, CFR 4.32%). In 2020, four regions were affected (Centre, Littoral, South, and South-West), with Littoral reporting the most cases (951 with 53 deaths, CFR 5.57%) and the Centre the fewest cases (63 cases without deaths, CFR 0%). In 2021, four regions were affected (Centre, Littoral, South, and South-West), with South-West reporting the most cases (556 with 21 deaths, CFR 3.78%) and Littoral the fewest cases (30 cases with 1 death, CFR 3.33%). The highest cholera burden during the study period was recorded in 2022, when eight regions were affected (Centre, East, Far-North, Littoral, North, West, South, and South-West), with Littoral reporting the most cases (7,093 with 153 deaths, CFR 2.16%) and East the fewest cases (12 cases with 2 deaths, CFR 16.67%). In 2023, six regions were affected (Centre, East, Littoral, West, South, and South-West), with the Centre reporting the most cases (4,653 with 155 deaths, CFR 3.33%) and East the fewest cases (8 cases with 3 deaths, CFR 37.50%).

Figure 1 illustrates the progression of cholera cases in Cameroon from 2016 to 2023, with red indicating districts which reported cases (Figure 1). The geographical distribution of cases fluctuates over the years, with recurring hotspots in the Littoral, Centre, and Far North regions. In 2016, cholera cases were primarily concentrated in the Littoral region and a few districts in the North. By 2017, there was a reduction in the number of cases, mainly affecting the Littoral and Far North regions. In 2018, there was a notable increase in cases, spreading across the Centre, Littoral, Far North, and West regions (Table 1, Figure 1).

**Figure 1:**
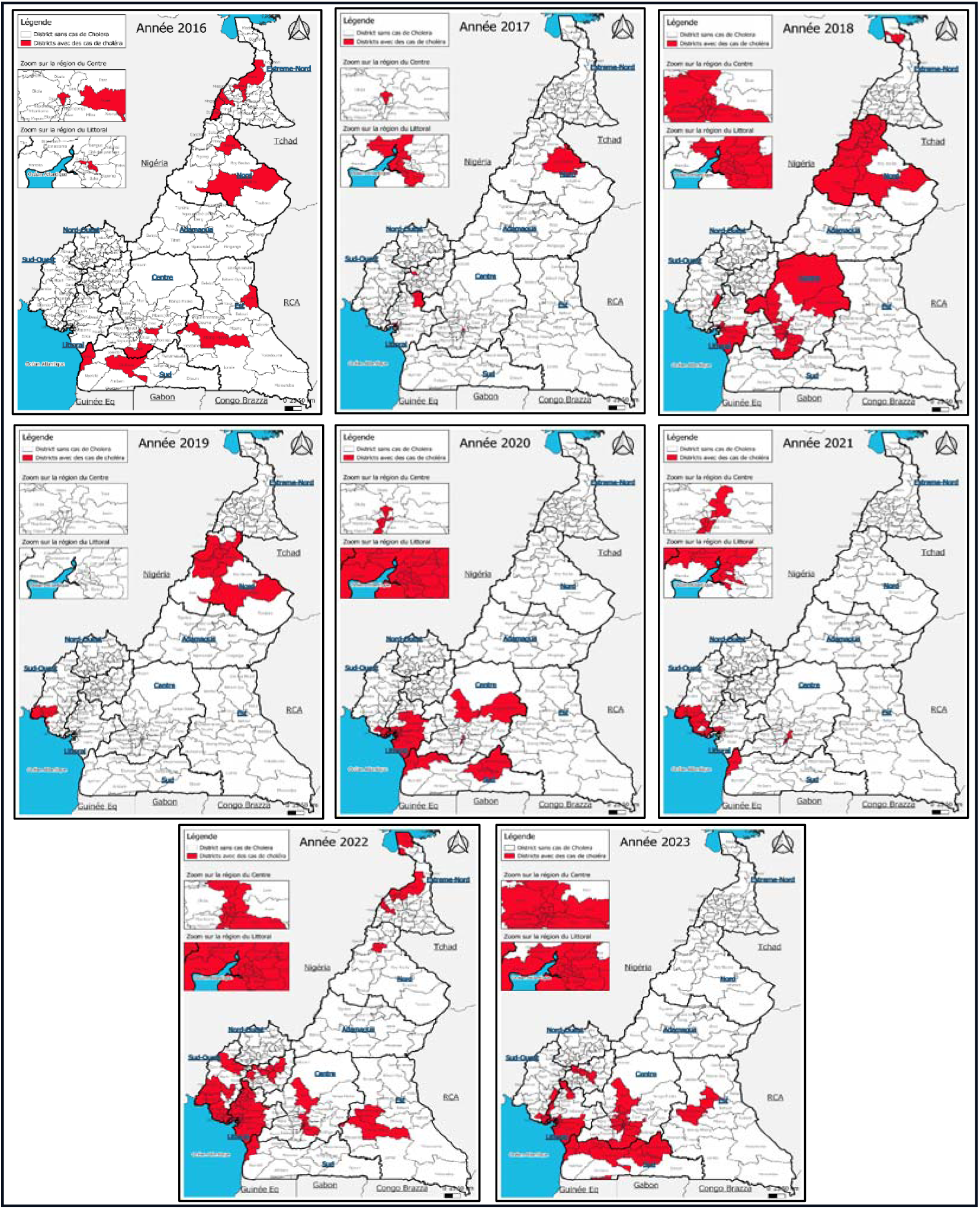
Mapping of cholera cases in Cameroon from 2016 to 2023

Between 2019 and 2021, there are fluctuating distributions of cases, but with persistent hotspots in the Littoral, Centre, and Far North regions. In 2022, there was a significant increase and broader dispersion of cases across these same regions. This trend continued in 2023, indicating ongoing cholera presence in these areas (Table 1, Figure 1).

### PAMIs identification using Priority Index and vulnerability

Nationwide, 35 health districts were identified with a priority index of at least 9. The Centre and Littoral regions alone account for more than 68% of the high-priority health districts, with 12 districts each (Tables 2 and 3).

**Table 2:**
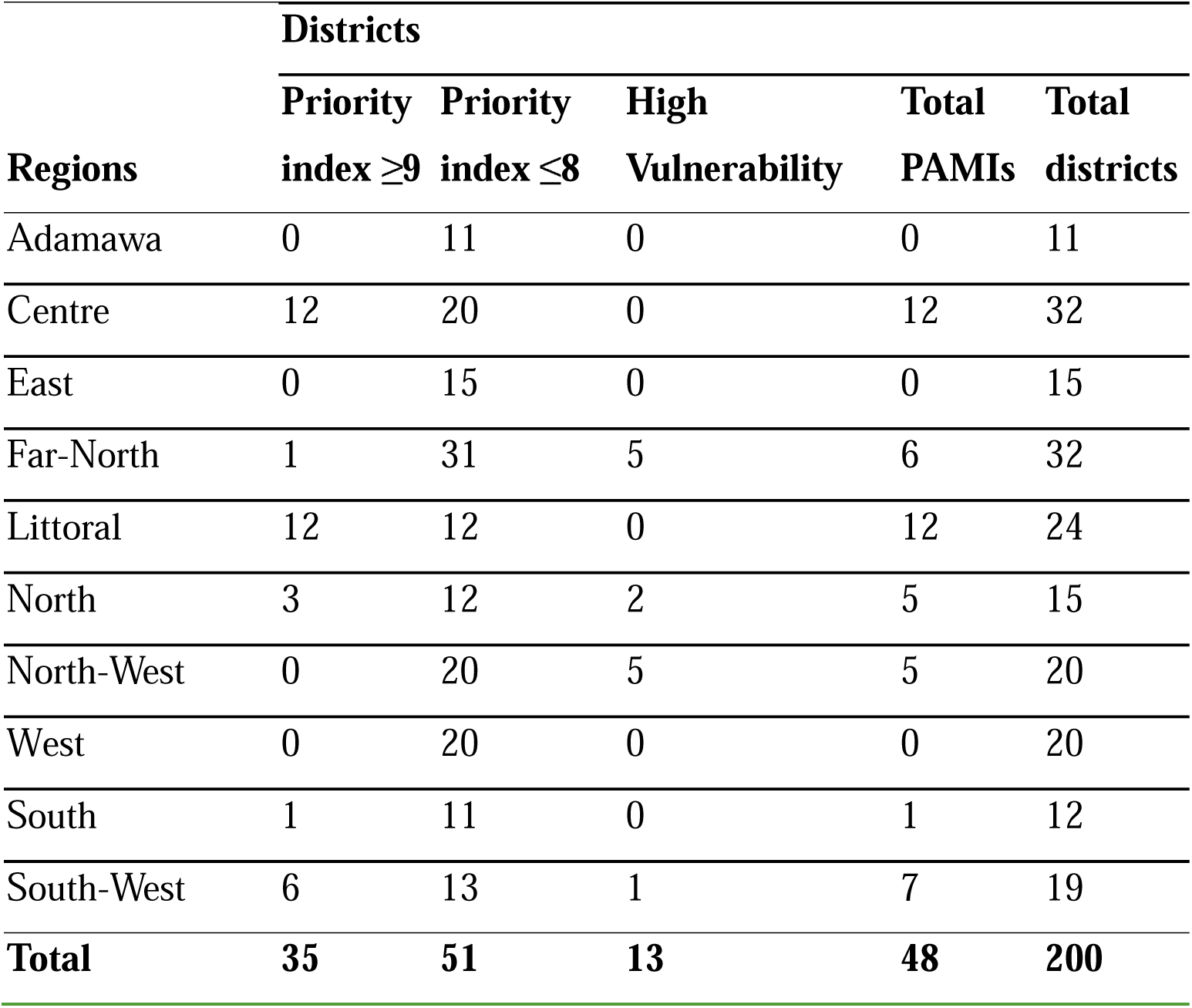
PAMIs identification based on priority index and vulnerability.

| Regions | Districts |  |  |  |  |
| --- | --- | --- | --- | --- | --- |
| | Priority index $\geq 9$ | Priority index $\leq 8$ | High Vulnerability | Total PAMIs | Total districts |
| Adamawa | 0 | 11 | 0 | 0 | 11 |
| Centre | 12 | 20 | 0 | 12 | 32 |
| East | 0 | 15 | 0 | 0 | 15 |
| Far-North | 1 | 31 | 5 | 6 | 32 |
| Littoral | 12 | 12 | 0 | 12 | 24 |
| North | 3 | 12 | 2 | 5 | 15 |
| North-West | 0 | 20 | 5 | 5 | 20 |
| West | 0 | 20 | 0 | 0 | 20 |
| South | 1 | 11 | 0 | 1 | 12 |
| South-West | 6 | 13 | 1 | 7 | 19 |
| **Total** | **35** | **51** | **13** | **48** | **200** |

**Table 3:** Final Priorization of PAMIs.

|  | **Region** | **Health Districts** | **Population size** | **Vulnerability index** | **Priority index using incidence, mortality, persistence, and positivity** |
| --- | --- | --- | --- | --- | --- |
| 1 | Centre | Biyem-Assi | 422270 | 5 | 10 |
| 2 | Centre | Cité-Verte | 472015 | 6 | 9 |
| 3 | Centre | Djoungolo | 575409 | 7 | 10 |
| 4 | Centre | Ebebda | 23602 | 5 | 9 |
| 5 | Centre | Efoulan | 480107 | 5 | 9 |
| 6 | Centre | Mfou | 112210 | 6 | 10 |
| 7 | Centre | Mvog-Ada | 411968 | 4 | 9 |
| 8 | Centre | Nkolbisson | 192440 | 6 | 10 |
| 9 | Centre | Nkolndongo | 599793 | 8 | 10 |
| 10 | Centre | Obala | 147550 | 4 | 12 |
| 11 | Centre | Odza | 442785 | 4 | 9 |
| 12 | Centre | Soa | 49369 | 5 | 10 |
| 13 | Far North | Makary | 144528 | 7 | 9 |
| 14 | Littoral | Bangue | 395315 | 9 | 10 |
| 15 | Littoral | Boko | 373983 | 7 | 11 |
| 16 | Littoral | Bonassama | 568369 | 6 | 11 |
| 17 | Littoral | Cité des palmiers | 348001 | 4 | 10 |
| 18 | Littoral | Deido | 640608 | 2 | 11 |
| 19 | Littoral | Japoma | 187006 | 9 | 10 |
| 20 | Littoral | Logbaba | 279923 | 4 | 9 |
| 21 | Littoral | Manoka | 31050 | 9 | 11 |
| 22 | Littoral | Melong | 112946 | 8 | 9 |
| 23 | Littoral | New-Bell | 324169 | 8 | 12 |
| 24 | Littoral | Njombe-Penja | 59250 | 8 | 11 |
| 25 | Littoral | Nylon | 462815 | 8 | 12 |
| 26 | North | Bibeni | 176023 | 7 | 9 |
| 27 | North | Golombe | 77146 | 5 | 9 |
| 28 | North | Pitoa | 172652 | 8 | 11 |
| 29 | South | Kribi | 118579 | 9 | 12 |
| 30 | South-West | Bakassi | 37390 | 7 | 11 |
| 31 | South-West | Buea | 188293 | 8 | 10 |
| 32 | South-West | Ekondo-Titi | 61031 | 8 | 11 |
| 33 | South-West | Limbé | 215410 | 9 | 11 |
| 34 | South-West | Mundemba | 23267 | 8 | 9 |
| 35 | South-West | Tiko | 164284 | 8 | 12 |
| 36 | Far North | Bourha | 90077 | 9 | 5 |
| 37 | Far North | Fotokol | 74704 | 10 | 7 |
| 38 | Far North | Mokolo | 333305 | 9 | 7 |
| 39 | Far North | Mora | 328959 | 9 | 6 |
| 40 | Far North | Yagoua | 272701 | 10 | 0 |
| 41 | North | Garoua I | 315119 | 9 | 8 |
| 42 | North | Touboro | 335091 | 9 | 0 |
| 43 | North-West | Ako | 66074 | 9 | 0 |
| 44 | North-West | Kumbo-East | 119735 | 9 | 0 |
| 45 | North-West | Kumbo-West | 108723 | 9 | 0 |
| 46 | North-West | Ndop | 193116 | 10 | 0 |
| 47 | North-West | Nwa | 60931 | 9 | 0 |
| 48 | South-West | Mbonge | 97999 | 10 | 0 |
| **Total population** |  |  | **11488090** |  |  |

Regarding vulnerability factors, health districts with priority indexes between 0 and 8 and exhibiting at least 9 vulnerability factors were selected as additional PAMIs (Table 2 and 3).

Regarding the final prioritization, Cameroon has set a threshold of 9 for both the priority index based on cholera incidence, mortality, persistence, and positivity, and using vulnerability factor index. In sum, based on the priority index, 35 priority health districts were identified as main PAMIs and 13 health districts were classified as additional PAMIs using the vulnerability index, yielding a total of 48 PAMIs in Cameroon for the study period (Table 3). The Centre and Littoral regions alone account for more than 50% of these PAMIs (Table 3).

Clustering of PAMIs is noticeable in 04 of 10 regions (Littoral, Centre, South-West, and the North) in Cameroon (Figure 2).

**Figure 2:**
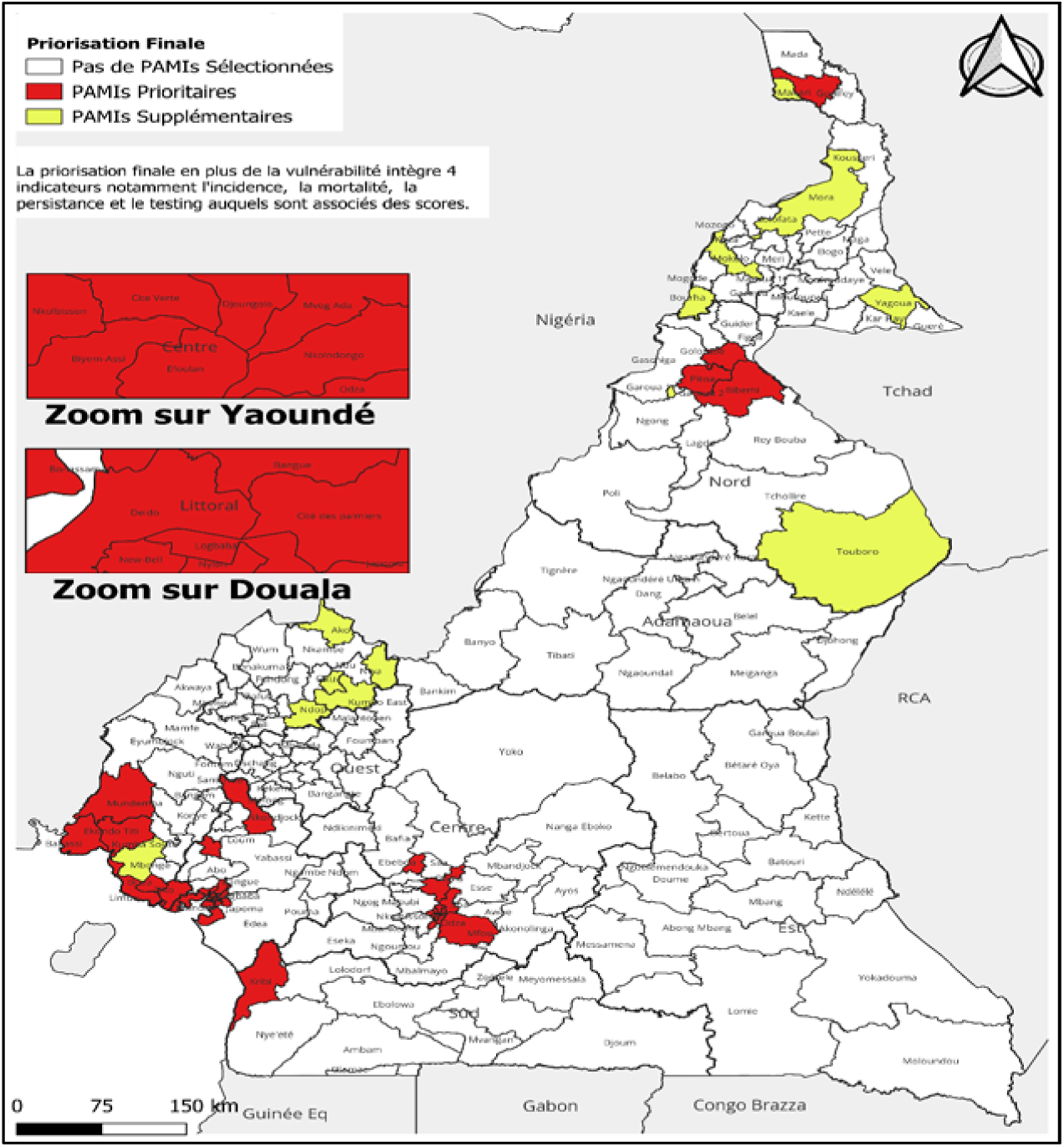
Mapping of PAMIs in Cameroon

## Discussion

This study highlights how historic epidemiological data and vulnerability factors can be used for prioritization of PAMIs. Using the GTFCC methodology, between 2016 and 2023, a total of 48 health districts in Cameroon were identified as PAMIs including 35 PAMIs based on priority index and 13 additional PAMIs based on vulnerability with an estimated average population at risk of 11 488 089, representing 42% of the total population of Cameroon. Our study emphasizes the urgent need to scale up sustainable cholera interventions in PAMIs in Cameroon to reverse this trend.

Over the last 8 years, 8 out of 10 regions recorded cholera cases in Cameroon. The longest and most intense cholera outbreak was recorded between 2021 and 2023, with 21,407 cases, including 516 deaths, yielding a high case fatality rate of 2.4%. Cholera epidemiology has been consistent in Cameroon over the study period affecting a minimum of 4 regions every year. However, between, the intensity of outbreaks was highest in the Northern regions of the country between 2018 and 2019, gradually reducing in 2020. In contrast, the high burden of cholera outbreaks increased in the Southern parts of the country since 2019. There was a high cholera persistence between 2018 and 2022 in the South-West region which was almost not affected by cholera outbreaks before 2018.

These trends in cholera outbreaks can be explained by factors including extreme climate situations (droughts and floods), a decreased intensity of the humanitarian crisis in the Far North and the worsened crisis in the Southwest and North-west regions leading to population displacement towards the Littoral, the West, and the Center regions. According to OCHA, Cameroon faces three complex and prolonged humanitarian crisis which increase population vulnerability. In February 2024, more than 1 million internally displaced people and nearly 500,000 refugees and asylum seekers were enumerated with 1.8 million people in need of life-saving access to water, hygiene and sanitation (WASH) in crisis-affected regions. In the Far North, as of April 2024, there were 573,263 people including IDPs, and refugees from Nigeria, due to violence and natural disasters. With only 40% of the population having access to improved WASH, the risk of cholera outbreak remains high in this part of the country. Violence and fear of attacks in the South-West and North-West regions lead to regular population displacements. Cameroon is also home to 353 000 refugees of the Central African Republic (CAR) who mostly live in the Eastern region of the country^19^. This volatile and unpredictable sociopolitical situation associated with extreme weather situations fuel cholera outbreaks. Indeed, a spatiotemporal analysis of cholera epidemics in Cameroon over the period of study is required. A similar dynamic pattern of PAMIs has been observed in Ethiopia^20^ which has been facing substantial humanitarian crisis associated with climate shocks, disease outbreaks, conflicts in a challenging socioeconomic context^21^. This calls for a regular update of the process of PAMI identification in such dynamic contexts, as recommended by the GTFCC^16^.

Half of all PAMIs is in urban areas of Center and Littoral regions which are home to the political and economic capitals, covering a population of 7 712 952 million and representing 28.5% of the country’s total population. Our results stress out population vulnerability in major urban areas of Cameroon and the resulting health insecurity. This might be due to internal migration, rapid urban growth, creation of informal shelters and insufficient town planning^22^. In Cameroon and elsewhere, migrants tend to live in slums^23^. In such informal settings, there is usually inadequate access to improved water hygiene and sanitation infrastructures as well as potable water, increasing the risk of cholera outbreaks^24^. In fact, there is tremendous shortage of water in urban and peri-urban areas in Cameroon^25^. Rapid urban growth also leads to limited access to healthcare, establishment of informal care structures. Cholera outbreaks happening in such contexts are associated with limited access to oral rehydration points for the most vulnerable and high cholera case fatality rates. In Cameroon, although there has been a decrease in cholera case fatality rates (CFR) over the study period, the overall CFR of 2.74% over the study period remains above the acceptable standard CFR of 1% which indicates early access to treatment^26,27^. These calls for the implementation of long-term targeted cholera interventions in PAMIs in Cameroon.

A proportion of 25% of all districts are at risk of cholera outbreaks with a total population 11 488 089 million population at risk of cholera representing 42% of the total population. As such Cameroon remains a country with high cholera transmission, implying that the country should engage in a control strategy in the national cholera plan rather than an elimination strategy. Similarly, countries like Burundi where PAMIs represent 25% of all health districts would opt for a cholera control strategy^28^. According to the GTFCC, to achieve cholera control or elimination, it is critical to organize activities around immunization, surveillance, risk communication and community engagement, WASH, health system strengthening, all with a good multisectoral coordination mechanism^14^ with a focus on PAMIs. The high number of people living at risk of cholera in PAMIs has financial consequences on the cholera control strategy in Cameroon. The use of OCV is recommended as a tool for long term cholera control through preventive immunization campaigns^29^. In Cameroon, this would mean targeting on average 20 million people living in 37 PAMIs which is a massive investment. Cameroon like several other countries has benefitted from financial support from GAVI for reactive immunization campaigns. Between 2021 and 2023, 8 million people have been immunized with OCV^13^. However, there has been a global shortage of OCV^30^ and this led to the initiation of a one-dose strategy for reactive campaigns in a context of high demand of OCV for reactive and preventive campaigns. This would likely lead to competitions between countries in need.

In addition to OCV use, there is need for substantial investments on surveillance systems and laboratory, WASH, community engagement and risk communication, health system strengthening, and most importantly coordination to sustain cholera control and reduce cholera mortality and morbidity in Cameroon. This would mean identifying targeted interventions around these pillars for implementation in each PAMIs, thereby requiring a massive investment. As such, it is critical for Cameroon to maintain the current cholera epidemiological status quo to gain time for resource mobilization for long term cholera control.

Despite filling an evidence gap for decision-making, our study does have limitations. The GTFCC’s methodology for PAMI identification considers geographic units as separate entities with their epidemiological data and vulnerability factors. It does not consider the risk of spread across neighboring districts. In addition, laboratory tests for cholera include rapid diagnostic tests and confirmation by culture or PCR in cholera reference laboratories. Laboratories are not available throughout the country, not always close to high-risk areas with ruptured reagents most of the time. Furthermore, transport media and RDTs are not always available in high-risk areas. All these can delay cholera case confirmation, transmission, outbreak intensity and duration. Nonetheless, according to the GTFCC guidelines for PAMI identification^17^, if testing indicators are deemed insufficient, positivity indicators should be removed. This was not the case in Cameroon. Furthermore, there was missing data for the period defined by the country in the newly created HDs (between 2019 and 2022). To limit bias and improve data reliability, a retrograde population estimate was made based on new list of health districts and the population growth rate in Cameroon. Finally, different data sources were used to calculate the priority index. As part of the process of this study, data curation for the defined period was carried out by the data management team to ensure quality and reliability of the data used. Nonetheless, this very first comprehensive analysis is an initial step for long term cholera control across Cameroon and beyond.

## Conclusion

In Cameroon, a total of 11,488,089 people living in 48 districts are at risk of cholera. This study sheds lights on the use of the GTFCC methodology to identify PAMIs in Cameroon. The use of epidemiological data and vulnerability data provide a better understanding of past cholera outbreaks, and this could serve as a basis for predicting future epidemics through modelling. In Cameroon, high-level commitment to the GTFCC roadmap to eliminate cholera by 2030 was followed by strong multisectoral, community and partner involvement in the process of developing the national cholera elimination plan with targeted interventions adapted to each PAMI. Overall, our findings are valuable evidence required for public health decision-making to level up cholera preparedness and prevention in Cameroon and Africa, ultimately contributing to a reduction in cholera deaths.

## Data Availability

All data produced in the present study are available upon reasonable request to the authors

## Acknowledgement

We would like to acknowledge all the Regional Delegations of Public Health, the Ministry of Public Health, the Ministry of Water, other sectoral administrations in charge of decentralization and regional development, external relations, institutional communication, youth and civic education, justice, higher school, secondary and primary education, finance, economy and planning, housing and urban development, environmental protection and sustainable development, defense and national security, and technical and financial Partners (WHO, CDC-Atlanta, UNICEF, USAID, IFCR, MSF, HEADA, MA Santé), civil society organisations (Cameroon Red Cross, French Red Cross, DEMTOU Humanitaire) and other agencies (the Centre Pasteur du Cameroun, CAMWATER, decentralized territorial authorities) for their support during this study and their commitment to the process of long-term cholera control.

